# Persistently high proportions of *Plasmodium*-infected *Anopheles funestus* mosquitoes in two villages in the Kilombero valley, south-eastern Tanzania

**DOI:** 10.1101/2021.06.24.21259445

**Authors:** Salum A. Mapua, Emmanuel E. Hape, Japhet Kihonda, Hamis Bwanary, Khamis Kifungo, Masoud Kilalangongono, Emmanuel W. Kaindoa, Halfan S. Ngowo, Fredros O. Okumu

**Author notes:** Corresponding author at: Department of Environmental Health and Ecological Sciences, Ifakara Health Institute.

## Abstract

**Background:** In south-eastern Tanzania where insecticide-treated nets have been widely used for more than 20 years, malaria transmission has greatly reduced but remains highly heterogenous over small distances. This study investigated the seasonal prevalence of *Plasmodium* sporozoite infections in the two main malaria vector species, *Anopheles funestus* and *Anopheles arabiensis* for 34 months, starting January 2018 to November 2020.

**Methods:** Adult mosquitoes were collected using CDC-light traps and Prokopack aspirators inside local houses in Igumbiro and Sululu villages, where earlier surveys had found very high densities of *An. funestus*. Collected females were sorted by taxa, and the samples examined using ELISA assays for detecting *Plasmodium* circumsporozoite protein in their salivary glands.

**Results:** Of 7,859 *An. funestus* tested, 4.6% (n = 365) were positive for *Pf* sporozoites in the salivary glands. On the contrary, only 0.4% (n = 9) of the 2,382 *An. arabiensis* tested were positive. The sporozoite prevalence did not vary significantly between the villages or seasons. Similarly, the proportions of parous females of either species were not significantly different between the two villages (p > 0.05) but was slightly higher in *An. funestus* (0.50) than in *An. arabiensis* (0.42). Analysis of the 2020 data determined that *An. funestus* contributed 97.7% of all malaria transmitted in households in these two villages.

**Conclusions:** In contexts where individual vector species mediate most of the pathogen transmission, it may be most appropriate to pursue a species-specific approach to better understand the ecology of the dominant vectors and target them with effective interventions to suppress transmission. Despite the widespread use and overall impact of insecticide-treated nets over the past two decades in the two study villages, there is still persistently high *Plasmodium* infection prevalence in local populations of *An. funestus*, which now carry ∼97% of all malaria infections and mediates intense year-round transmission. Further reduction in malaria burden in these or other similar settings requires effective targeting of *An. funestus*.

## 1. Introduction

Starting two decades ago when more than a million lives were being lost to malaria yearly [1], there have been significant gains, including ∼60% reduction in deaths, despite doubling of Africa’s population [2]. Insecticide-treated nets (ITNs), indoor residual spraying (IRS) and effective case management have contributed greatly to these gains [3,4]. The persisting malaria transmission in most endemic settings is now thought to be largely due to widespread insecticide resistance among major malaria vectors [5–8], behavioral adaptation of malaria vectors to indoor interventions [9–11], parasite resistance to anti-malarial drugs [12,13], certain human behaviors and activities [14–16] and asymptomatic infections in older age groups such as school-aged children [17]. Other factors such as inadequate environmental sanitation, poor housing conditions and low household incomes may also be perpetuating risk in many endemic villages [18,19], despite more than two decades of widespread ITN use.

In rural south-eastern Tanzanian villages, where ITNs have been widely used for more than 20 years, malaria transmission intensities have greatly reduced, in some cases from highs of >300 infectious bites per person per year (ib/p/y) in early 2000s to new averages below 30 ib/p/y [20]. These gains have been mostly attributed to widespread use of ITNs and effective case management [20,21]. Today, malaria transmission remains highly varied over small distances [20,22], and recent parasite surveys have demonstrated ranges of <1% prevalence in urban centers of Ifakara town (<150m above sea level) to highs of >40% in some higher altitude villages (∼300m above sea level) just 30km away (Swai *et al*., unpublished).

Recent studies in Ulanga and Kilombero districts, have demonstrated that *Anopheles funestus* alone now contributes nearly nine in every ten new infections, even in villages where it is outnumbered by *Anopheles arabiensis* [6,23]. The species is also strongly resistant to pyrethroids used in the ITNs [24], survives much longer than its counterparts [6,25] and is strongly adapted to primarily bite humans over other vertebrates [26]. Historically, *Anopheles gambiae* sensu stricto dominated transmission in Ulanga and Kilombero districts, with the *An. funestus* being another important vector during the dry season [27]. However, *An. gambiae* s.s population and contribution to the malaria transmission in these districts started to decline following introduction of the ITNs [22,23], and reached undetectable level by 2012 [23]. Recent studies have confirmed absence of the *An. gambiae* s.s in these districts, leaving the sibling species *An. arabiensis* at large [6,18,28,29]. While no detailed analysis has been conducted, it is hypothesized that villages currently having the greatest malaria prevalence are those with highest densities of *An. funestus*. Moreover, given its preference for permanent and semi-permanent aquatic habitats which last far longer than the rainy season [30], this species is thought to be important for year-long transmission of malaria.

This current analysis was conducted in two villages, previously identified as having high densities of *An. funestus* [31], and coincidentally having very high malaria prevalence estimates (Swai *et al*., unpublished). The aim was to analyze and compare seasonal prevalence of *Plasmodium* sporozoite infections in the main malaria vectors, *An. funestus* and *An. arabiensis* following nearly two decades of widespread ITN use.

## 2. Methods

### 2.1 Study area

The study was done in Ulanga and Kilombero districts, in the Kilombero valley in south-eastern Tanzania (Figure 1). Mosquitoes were collected from two villages namely Sululu in Kilombero district (8.00324°S, 36.83118°E) and Igumbiro in Ulanga district (8.35021°S, 36.67299°E). Average annual rainfall was 1200 – 1600 mm and mean annual temperatures were 20 – 32°C [28]. Most residents here are subsistence farmers, cultivating mainly rice but also other crops such as maize, beans and sweet potatoes [32]. According to 2012 national population census, Igumbiro and Sululu villages are within wards with the population of 16,329 and 9,048 respectively [33]. Sululu village sits on higher altitude above sea level compared to Igumbiro village (Figure 1).

**Figure 1:** Map showing villages in the Kilombero valley, south-eastern Tanzania where adult *Anopheles funestus* and *Anopheles arabiensis* mosquitoes were collected.

### 2.2 Mosquito collection and processing

Mosquitoes were routinely collected from January 2018 to December 2020. In each village, sampling was done in 10-20 houses for five nights each week from January 2018 to January 2020, as part of the ongoing project that focused mainly on *An. funestus* mosquitoes. However, from March 2020 only three households were sampled nightly for four nights each week to December 2020, as part of another project focused on both *An. funestus* and *An. arabiensis* mosquitoes. Herein, data from both aforementioned projects were combined and analysed. All collections were done indoors, CDC light traps [34] and Prokopack aspirators [35] were used to sample host-seeking and resting mosquitoes respectively. Female mosquitoes were identified and sorted by taxa and physiological state [36], after which *An. arabiensis and An. funestus* were packed in batches of ten mosquitoes in 1.5 ml microcentrifuge tubes for circumsporozoite enzyme-linked immunosorbent assays (ELISA) [37], of which only a pool of heads and thoraces were used. The ELISA lysates were boiled for 10 minutes at 100 °C to eliminate heat-labile non *Plasmodium* protozoan antigens that will render false positivity [38]. A sub-sample of the mosquitoes collected from March 2020 to December 2020 were subjected to dissection to assess parity status as described by Kaindoa *et al* [6]. Recent studies in the area had determined that indoor collections of *An. gambiae* s.l. consisted only of *An. arabiensis* (100%), and those of *An. funestus* group were mainly *An. funestus* s.s [6,28].

### 2.3 Statistical analysis

R statistical software version 3.6.0 [39] was used to execute a simple generalized linear model (GLM) with binomial distributions to examine odds of sporozoite-infected *Anopheles* and also parous mosquitoes. Generalized linear mixed models (GLMM) were used to model the abundance of different mosquito species. Here the number of mosquitoes caught were modelled as a response variable using a negative binomial variate to account for the over-dispersion in the number of catches. Sampling location and season were included as the fixed terms. To account for the unexplained variation between sampling days, sampling dates nested within a month of the collection was included as random term. Relative risk and their corresponding 95% CI were reported. Entomological inoculation rate (EIR) was estimated as a function of a biting rate and a proportion of mosquitoes tested positive with *Plasmodium* sporozoite. The annual EIR is estimated by multiplying a daily EIR with 365 (Annual EIR = daily biting rate ⍰ sporozoite rate ⍰ 365). The adjusted annual EIR was estimated by the same function except that, the corrected biting rate was used in the place of daily biting rate as described by Kaindoa *et al* [6]. Herein, the dry season comprises of months from January to May, and from June to December for the rainy season.

## 3. Results

A total of 306,589 mosquitoes were collected with 53.8% (n = 165,058) being *An. funestus* mosquitoes. A sub-sample of the collected mosquitoes (n = 10,241) were analyzed for *Plasmodium* sporozoite infections, of which 23.3% (n = 2,382) were *An. arabiensis* and 76.7% (n = 7859), *An. funestus*. Of 7,859 *An. funes*tus tested, 4.6% (n = 365) were positive for *Plasmodium* sporozoites in the salivary glands. On the contrary, only 0.4% (n = 9) of the 2,382 *An. arabiensis* tested were positive. Though the sporozoite prevalence in Sululu appeared slightly higher than in Igumbiro village (Table 1), this analysis showed no statistically significant difference in the sporozoite prevalence for *An. funestus* between villages or seasons (p > 0.05). The *An. arabiensis* mosquitoes however had higher sporozoite prevalence in Sululu than in Igumbiro village (Table 1).

**Table 1:** Results of the multivariate analysis of *Plasmodium falciparum* sporozoites infectivity in *Anopheles funestus* and *Anopheles arabiensis* mosquitoes by village and season

| Species | Variable |  | Total (N) | Sporozoite prevalence<br>n (%) | OR (95% LC-UC) | p-value |
| --- | --- | --- | --- | --- | --- | --- |
| *Anopheles funestus* | Season | Dry | 4425 | 199 (4.5) | 1 | 0.54 |
|  |  | Rainy | 3434 | 166 (4.8) | 1.07 (0.86-1.32) |  |
|  | Village | Igumbiro | 3914 | 175 (4.5) | 1 | 0.56 |
|  |  | Sululu | 3945 | 190 (4.8) | 1.07 (0.86-1.32) |  |
| *Anopheles arabiensis* | Season | Dry | 2172 | 8 (0.4) | 1 | 0.95 |
|  |  | Rainy | 210 | 1 (0.5) | 0.93 (0.11-7.70) |  |
|  | Village | Igumbiro | 1851 | 4 (0.2) | 1 | 0.03 |
|  |  | Sululu | 531 | 5 (0.9) | 4.41 (1.17-16.69) |  |

\*Percentage sporozoite-prevalence = Sporozoite positive (n)/Total number of mosquitoes analysed (N)

The parity dissections determined that 50.3% of the *An. funestus* (n/N = 80/159) and 41.8% of the *An. arabiensis females* (n/N = 66/158) were parous (Table 2). There was no statistical difference in parity between villages for either species (Table 2). Proportion of *An. funestus* positive for *Pf* sporozoite was high in both seasons (Table 1), and both villages recorded high densities of both vector species throughout the year (Table 3). Analysis of the year-to-year data showed that in 2020, 97.7% of all malaria transmission events were mediated by *An. funestus*, and that *An. arabiensis* played only a minor role (Table 4).

**Table 2:** Results of the multivariate analysis of parity in *Anopheles funestus* and *Anopheles arabiensis* mosquitoes by village Percentage parous = parous (n)/Total number of mosquitoes examined (N)

| Species | Village | Total (N) | Parous n (%) | OR (95% LC-UC) | p-value |
| --- | --- | --- | --- | --- | --- |
| *Anopheles funestus* | Igumbiro | 79 | 39 (49.4) | 1 | 0.81 |
|  | Sululu | 80 | 41 (51.3) | 1.08 (0.58-2.01) |  |
| *Anopheles arabiensis* | Igumbiro | 78 | 35 (44.9) | 1 | 0.44 |
|  | Sululu | 80 | 31 (38.8) | 0.78 (0.41-1.46) |  |

\*Percentage parous = parous (n)/Total number of mosquitoes examined (N)

**Table 3:** Results of the multivariate analysis of biting densities of *Anopheles funestus* and *Anopheles arabiensis* mosquitoes by village and season

| Species | Village/Season | | Total | Mean ± 2SE | RR (95% LC-UC) | p-value |
| --- | --- | --- | --- | --- | --- | --- |
| *Anopheles arabiensis* | Village | Igumbiro | 88149 | 18 ± 0.5 | 1 | <0.001 |
| | | Sululu | 51000 | 6.9 ± 0.3 | 0.26 (0.25-0.28) | |
| | Season | Dry | 74819 | 9.5 ± 0.4 | 1 | <0.001 |
| | | Rainy | 64330 | 14.4 ± 0.5 | 2.54 (1.74-3.72) | |
| *Anopheles funestus* | Village | Igumbiro | 55617 | 11.4 ± 0.3 | 1 | 0.492 |
| | | Sululu | 101582 | 13.7 ± 0.5 | 1.02 (0.97-1.07) | |
| | Season | Dry | 83475 | 10.6 ± 0.3 | 1 | 0.374 |
| | | Rainy | 73724 | 16.5 ± 0.6 | 1.26 (0.76-2.07) | |

**Table 4:**
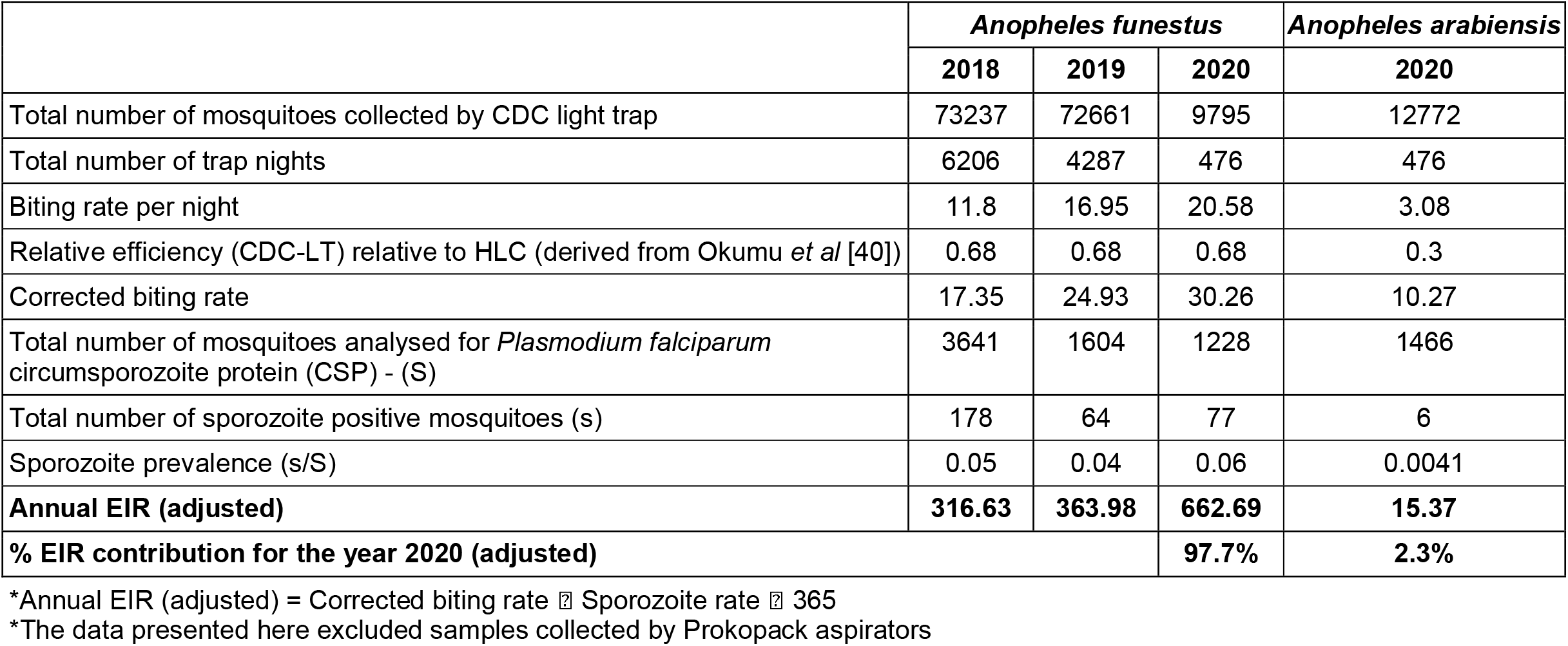
Infectious status of *Anopheles funestus* and *Anopheles arabiensis* mosquitoes collected from 2018 to 2020

**Table 5:**
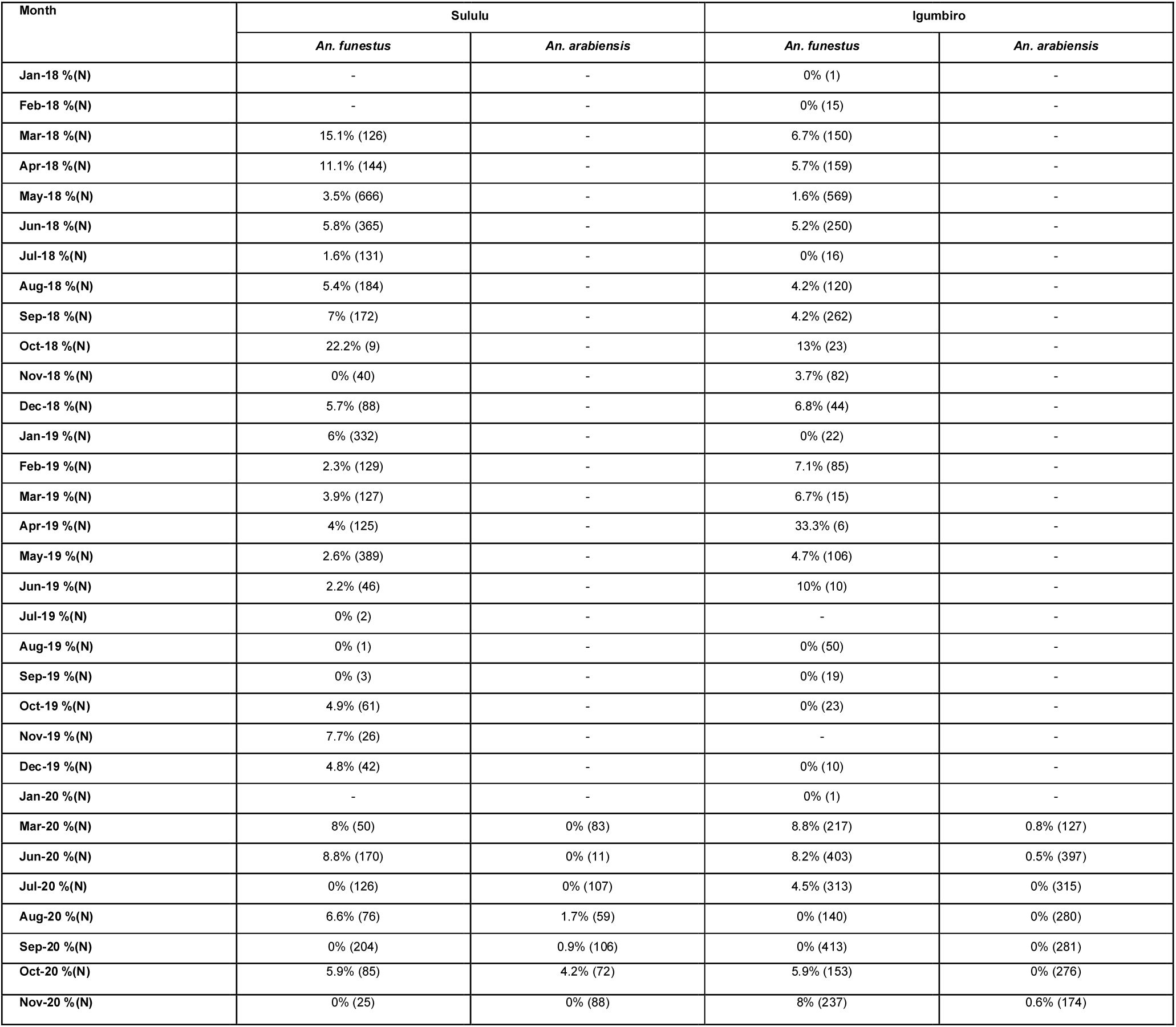
*Plasmodium* sporozoite infectivity by village, mosquito species and month.

## 4. Discussion

Recent evidence has shown that *An. funestus* now mediates most of the ongoing malaria transmission in many countries in East and Southern Africa [6,41,42]. In rural south-eastern Tanzania, this species carries more than 85% of all malaria infections even in villages where it occurs at lower frequencies than *An. arabiensis* [6,21,43]. malaria transmission has declined significantly in the Kilombero valley since 2000, mostly due to the wide coverage of ITNs and effective case management [20,22].

This short report presents an analysis of mosquitoes collected between 2018 and 2020 in two villages previously identified as having very high densities *of An. funestus*. It had been hypothesized that the proven dominance of *An. funestus* in the valley, coupled with the high densities of this vector species, as well as its strong pyrethroid-resistance status and greater survival in nature may lead to high residual transmission burden in the relevant villages, and that this situation may make malaria transmission control much more difficult than elsewhere. This analysis confirms the high parity rates but further presents two surprising findings. First is the extremely high *Plasmodium* infection prevalence in the *An. funestus* mosquitoes over the three years of sampling, averaging 4.6% despite high ITN use in the areas. Given the nightly mosquito catches by CDC light traps, this translates to 447.8 infectious bites/person/year, which is considerably higher than most recent estimates [6,21]. The second finding was that the high infection prevalence rates were consistent throughout dry and rainy season for the entire duration of the study. This suggests that *An. funestus* not only plays an important role in transmission but also that it mediates transmission throughout the whole year. There were months with very high transmission and months with very low transmission, but when data was aggregated by season, there was no significant difference in transmission intensities. Additional studies will be necessary to understand the dry season ecology *of An. funestus* and the factors driving transmission at different times of the year, so as to design effective tools towards reducing malaria transmission. Additionally, the higher sporozoite prevalence observed in *An. arabiensis* in one of the village (i.e. Sululu) corroborate recent finding (Swai *et al*., unpublished) that indicate geographical heterogeneity in transmission over small distances. Thus, understanding local vectors’ ecology is crucial for control measures. Polymerase chain reaction (PCR) may have greater sensitivity in detection of *Plasmodium* sporozoite, however, these improvements are marginal and practically irrelevant in areas with moderate to high transmission such as south-eastern Tanzanian districts of Ulanga and Kilombero.

Kaindoa *et al*. [6] reported *An. funestus* mosquitoes carrying 86% of malaria infections in Kilombero valley. Lwetoijera *et al*. also demonstrated significant role of *An. funestus* following the decline of *An. gambiae*, formerly the most important vector in the valley [23]. Since then, several other studies have confirmed the dominance of *An. funestus* in this area [20]. In addition, reports of higher entomological inoculation rates by *An. funestus* have also been documented elsewhere [44–46], however, the rates were not as high. Other studies have also shown the strong pyrethroid resistance, and that while its aquatic ecology is still poorly understood, it occupies perennial habitats which remain water-filled most of the year [30]. This ecological characteristic may explain its dominance in densities and transmission activity throughout the year. It is clear therefore that efforts to further reduce malaria transmission by this vector must consider specific measures targeting *An. funestus* so as to strongly diminish its potential.

Opportunities for improved control of *An. funestus* in the area may include new generation ITNs with synergists or multiple actives [47–50], or use of larval source management, which would be effective even against the pyrethroid-resistant populations. Fortunately, the current National Malaria Strategic Plan (NMSP) of Tanzania encourage implementation of larviciding in rural settings [51]. Coupled with recent findings of Nambunga *et al*. [30] which indicate that aquatic habitats of *An. funestus* mosquitoes in Kilombero valley falls within few, fixed and findable criterion of World Health Organization [52]. Thus, provide possibilities of a cost-effective and plausible species-specific intervention to significantly reduce malaria transmission in this valley. Additionally, susceptibility of *An. funestus* towards organophosphate notably pirimiphos methyl poses a potential opportunity for control. Pirimiphos methyl is already approved and used for IRS in Tanzania [53,54], thus there is a need to design cost-effective tools that may exploit the possibility of using pirimiphos methyl.

## 5. Conclusion

This study demonstrates that despite the widespread use and overall impact of ITNs, there is still persistently high *Plasmodium* infection prevalence in the dominant malaria vector, *An. funestus*, causing intense year-round malaria transmission in the study villages. Further reduction in malaria burden in this and similar settings thus requires effective targeting of *An. funestus*. The study also demonstrates that in certain contexts such as these, where one species is mediating most of the pathogen transmission, there could be significant potential in pursuing a species-specific approach for vector control by investigating and targeting the dominant vector species to suppress transmission.

## Data Availability

Data used to generate our findings can be accessed upon reasonable request to the corresponding author.

## Abbreviations

CDC: Centers for Diseases Control and Prevention
LLINs: Long-Lasting Insecticidal Nets
NMSP: National Malaria Strategic Plan
ITN: Insecticide-Treated Net
IRS: Indoor Residual Spraying
ELISA: Enzyme-Linked Immunosorbent Assay
GLM: Generalized Linear Model
GLMM: Generalized Linear Mixed Model
*Pf*-CSP: *Plasmodium falciparum* Circumsporozoite Protein
PCR: Polymerase Chain Reaction

## Declarations

### Ethical approval and consent to participate

Ethical approvals for this project were obtained from Ifakara Health Institute’s Institutional Review Board (Ref. IHI/IRB/No: 007 - 2018) and the Medical Research Coordinating Committee (MRCC) at the National Institute for Medical Research, in Tanzania (Ref: NIMR/HQ/R.8a/Vol. IX/2895). Written consents were sought from all participants of this study, after they had understood the purpose and procedure of the discussions.

### Consent for publication

Permission to publish this study was obtained from National institute for Medical Research, in Tanzania (NIMR/HQ/P.12 VOL XXXII/144).

### Authors contributions

SAM, FOO, HSN and EEH were involved in study design. SAM, EEH, HSN, JK, HB, KK and MK were involved in data collection. SAM and HSN conducted data analysis. SAM wrote the manuscript. FOO, HSN, EWK and EEH provided thorough review of the manuscript. All authors read and approved the final manuscript.

## Acknowledgments

We would like to thank village leaders and community members in Igumbiro and Sululu Villages for allowing their houses and areas to be used during mosquito collection. We also extend our gratitude to Joseph Mgando, Neema Nombo, Dickson Mwasheshi and Godfrey Matanila for their assistance during mosquito samples’ processing. We are also grateful to Najat Kahamba, Doreen Siria, Rukiyah Njalambaha, Augusto Mwambaluka, Jonael Msangi, Said Abbasi, Swaleh Masoud and Francis Tumbo for their valuable assistance in making this work feasible.

## Reference

1. World Health Organization. WHO expert committee on malaria [Internet]. World Health Organization - Technical Report Series. 2000. Available: https://apps.who.int/iris/handle/10665/42247

2. World Health Organization. World Malaria Report 2019. Geneva. [Internet]. 2019. Available: https://www.who.int/publications-detail/world-malaria-report-2019

3. Bhatt S, Weiss DJ, Cameron E, Bisanzio D, Mappin B, Dalrymple U, et al. The effect of malaria control on Plasmodium falciparum in Africa between 2000 and 2015. Nature. 2015; doi:10.1038/nature15535

4. Steketee, Richard W. and CCC. Impact of national malaria control scale-up programmes in Africa: magnitude and attribution of effects. Malar J. 2010;299.

5. Matowo NS, Munhenga G, Tanner M, Coetzee M, Feringa WF, Ngowo HS, et al. Fine-scale spatial and temporal heterogeneities in insecticide resistance profiles of the malaria vector, Anopheles arabiensis in rural south-eastern Tanzania [version 1; referees: 2 approved]. Wellcome Open Res. 2017; doi:10.12688/wellcomeopenres.12617.1

6. Kaindoa EW, Matowo NS, Ngowo HS, Mkandawile G, Mmbando A, Finda M, et al. Interventions that effectively target Anopheles funestus mosquitoes could significantly improve control of persistent malaria transmission in south-eastern Tanzania. PLoS One. 2017;12. doi:10.1371/journal.pone.0177807

7. Casimiro S, Coleman M, Mohloai P, Hemingway J, Sharp B. Insecticide resistance in Anopheles funestus (Diptera: Culicidae) from Mozambique. J Med Entomol. 2006; doi:10.1603/0022-2585(2006)043[0267:IRIAFD]2.0.CO;2

8. Cuamba N, Morgan JC, Irving H, Steven A, Wondji CS. High level of pyrethroid resistance in an Anopheles funestus population of the chokwe district inmozambique. PLoS One. 2010;5. doi:10.1371/journal.pone.0011010

9. Russell TL, Govella NJ, Azizi S, Drakeley CJ, Kachur SP, Killeen GF. Increased proportions of outdoor feeding among residual malaria vector populations following increased use of insecticide-treated nets in rural Tanzania. Malar J. 2011;10. doi:10.1186/1475-2875-10-80

10. Sougoufara S, Diédhiou SM, Doucouré S, Diagne N, Sembène PM, Harry M, et al. Biting by Anopheles funestus in broad daylight after use of long-lasting insecticidal nets: A new challenge to malaria elimination. Malar J. 2014; doi:10.1186/1475-2875-13-125

11. Moiroux N, Gomez MB, Pennetier C, Elanga E, Djènontin A, Chandre F, et al. Changes in anopheles funestus biting behavior following universal coverage of long-lasting insecticidal nets in benin. J Infect Dis. 2012; doi:10.1093/infdis/jis565

12. Ashley EA, Dhorda M, Fairhurst RM, Amaratunga C, Lim P, Suon S, et al. Spread of artemisinin resistance in Plasmodium falciparum malaria. N Engl J Med. 2014; doi:10.1056/NEJMoa1314981

13. Dondorp AM, Yeung S, White L, Nguon C, Day NPJ, Socheat D, et al. Artemisinin resistance: Current status and scenarios for containment. Nature Reviews Microbiology. 2010. doi:10.1038/nrmicro2331

14. Matowo NS, Moore J, Mapua S, Madumla EP, Moshi IR, Kaindoa EW, et al. Using a new odour-baited device to explore options for luring and killing outdoor-biting malaria vectors: A report on design and field evaluation of the Mosquito Landing Box. Parasites and Vectors. 2013;6. doi:10.1186/1756-3305-6-137

15. Finda MF, Moshi IR, Monroe A, Limwagu AJ, Nyoni AP, Swai JK, et al. Linking human behaviours and malaria vector biting risk in south-eastern Tanzania. PLoS One. 2019; doi:10.1371/journal.pone.0217414

16. Monroe A, Monroe A, Monroe A, Moore S, Moore S, Moore S, et al. Methods and indicators for measuring patterns of human exposure to malaria vectors. Malar J. 2020; doi:10.1186/s12936-020-03271-z

17. Andolina C, Rek JC, Briggs J, Okoth J, Musiime A, Ramjith J, et al. Sources of persistent malaria transmission in a setting with e ff ective malaria control in eastern Uganda_l_: a longitudinal, observational cohort study. 2021;3099.

18. Kaindoa EW, Finda M, Kiplagat J, Mkandawile G, Nyoni A, Coetzee M, et al. Housing gaps, mosquitoes and public viewpoints: A mixed methods assessment of relationships between house characteristics, malaria vector biting risk and community perspectives in rural Tanzania. Malar J. 2018; doi:10.1186/s12936-018-2450-y

19. Liu JX, Bousema T, Zelman B, Gesase S, Hashim R, Maxwell C, et al. Is housing quality associated with malaria incidence among young children and mosquito vector numbers? Evidence from Korogwe, Tanzania. PLoS One. 2014;9. doi:10.1371/journal.pone.0087358

20. Okumu F, Finda M. Key Characteristics of Residual Malaria Transmission in Two Districts in South-Eastern Tanzania—Implications for Improved Control. J Infect Dis. 2021;223: 15–17. doi:10.1093/infdis/jiaa653

21. Finda MF, Limwagu AJ, Ngowo HS, Matowo NS, Swai JK, Kaindoa E, et al. Dramatic decreases of malaria transmission intensities in Ifakara, south-eastern Tanzania since early 2000s. Malar J. 2018; doi:10.1186/s12936-018-2511-2

22. Russell TL, Lwetoijera DW, Maliti D, Chipwaza B, Kihonda J, Charlwood JD, et al. Impact of promoting longer-lasting insecticide treatment of bed nets upon malaria transmission in a rural Tanzanian setting with pre-existing high coverage of untreated nets. Malar J. 2010;9. doi:10.1186/1475-2875-9-187

23. Lwetoijera DW, Harris C, Kiware SS, Dongus S, Devine GJ, McCall PJ, et al. Increasing role of Anopheles funestus and Anopheles arabiensis in malaria transmission in the Kilombero Valley, Tanzania. Malar J. 2014;13. doi:10.1186/1475-2875-13-331

24. Pinda PG, Eichenberger C, Ngowo HS, Msaky DS, Abbasi S, Kihonda J, et al. Comparative assessment of insecticide resistance phenotypes in two major malaria vectors, Anopheles funestus and Anopheles arabiensis in south-eastern Tanzania. Malar J. 2020; doi:10.1186/s12936-020-03483-3

25. Ngowo HS, Hape EE, Matthiopoulos J, Ferguson HM, Okumu FO. Fitness characteristics of the malaria vector Anopheles funestus during an attempted laboratory colonization. Malar J. 2021;20. doi:10.1186/s12936-021-03677-3

26. Takken W, Verhulst NO. Host preferences of blood-feeding mosquitoes. Annual Review of Entomology. 2013. doi:10.1146/annurev-ento-120811-153618

27. Charlwood JD, Vij R, Billingsley PF. Dry season refugia of malaria-transmitting mosquitoes in a dry savannah zone of east Africa. Am J Trop Med Hyg. 2000;62. doi:10.4269/ajtmh.2000.62.726

28. Ngowo, H. S., Kaindoa, E. W., Matthiopoulos, J., Ferguson, H. M., & Okumu FO. Variations in household microclimate affect outdoor-biting behaviour of malaria vectors. Wellcome open Res. 2017;2.

29. Limwagu AJ, Kaindoa EW, Ngowo HS, Hape E, Finda M, Mkandawile G, et al. Using a miniaturized double-net trap (DN-Mini) to assess relationships between indoor-outdoor biting preferences and physiological ages of two malaria vectors, Anopheles arabiensis and Anopheles funestus. Malar J. 2019; doi:10.1186/s12936-019-2913-9

30. Nambunga IH, Ngowo HS, Mapua SA, Hape EE, Msugupakulya BJ, Msaky DS, et al. Aquatic habitats of the malaria vector Anopheles funestus in rural south-eastern Tanzania. Malar J. 2020; doi:10.1186/s12936-020-03295-5

31. Kaindoa EW, Ngowo HS, Limwagu AJ, Tchouakui M, Hape E, Abbasi S, et al. Swarms of the malaria vector Anopheles funestus in Tanzania. Malar J. 2019; doi:10.1186/s12936-019-2660-y

32. Swai JK, Finda MF, Madumla EP, Lingamba GF, Moshi IR, Rafiq MY, et al. Studies on mosquito biting risk among migratory rice farmers in rural south-eastern Tanzania and development of a portable mosquito-proof hut. Malar J. 2016; doi:10.1186/s12936-016-1616-8

33. National Bureau of Statistics. 2012 population and housing census, population distribution by administrative areas. 2013.

34. Mboera LEG, Kihonda J, Braks MAH, Knols BGJ. Short report: Influence of centers for disease control light trap position, relative to a human-baited bed net, on catches of Anopheles gambiae and Culex quinquefasciatus in Tanzania. Am J Trop Med Hyg. 1998;59: 595–596. doi:10.4269/ajtmh.1998.59.595

35. Maia MF, Robinson A, John A, Mgando J, Simfukwe E, Moore SJ. Comparison of the CDC Backpack aspirator and the Prokopack aspirator for sampling indoor-and outdoor-resting mosquitoes in southern Tanzania. Parasites and Vectors. 2011;4. doi:10.1186/1756-3305-4-124

36. Gillies MT, De Meillon B. The Anophelinae of Africa south of the Sahara (Ethiopian zoogeographical region). Sahara Ethiop Zoogeographical. 1968;

37. Beier JC, Perkins P V., Koros JK, Onyango FK, Gargan TP, Wirtz RA, et al. Malaria sporozoite detection by dissection and ELISA to assess infectivity of afrotropical Anopheles (Diptera: Culicidae). J Med Entomol. 1990; doi:10.1093/jmedent/27.3.377

38. Durnez L, Van Bortel W, Denis L, Roelants P, Veracx A, Trung HD, et al. False positive circumsporozoite protein ELISA: A challenge for the estimation of the entomological inoculation rate of malaria and for vector incrimination. Malar J. 2011; doi:10.1186/1475-2875-10-195

39. R Development Core Team R. R: A Language and Environment for Statistical Computing [Internet]. R Foundation for Statistical Computing. 2019. p. 409. doi:10.1007/978-3-540-74686-7

40. Okumu FO, Kotas ME, Kihonda J, Mathenge E, Killeen GF, Moore SJ. Comparative Evaluation of Methods Used for Sampling Malaria Vectors in the Kilombero Valley, South Eastern Tanzania. Open Trop Med J. 2008; doi:10.2174/1874315300801010051

41. Burke A, Dahan-Moss Y, Duncan F, Qwabe B, Coetzee M, Koekemoer L, et al. Anopheles parensis contributes to residual malaria transmission in South Africa. Malar J. 2019; doi:10.1186/s12936-019-2889-5

42. McCann RS, Ochomo E, Bayoh MN, Vulule JM, Hamel MJ, Gimnig JE, et al. Reemergence of Anopheles funestus as a vector of Plasmodium falciparum in Western Kenya after long-term implementation of insecticide-treated bed nets. Am J Trop Med Hyg. 2014; doi:10.4269/ajtmh.13-0614

43. Swai JK, Mmbando AS, Ngowo HS, Odufuwa OG, Finda MF, Mponzi W, et al. Protecting migratory farmers in rural Tanzania using eave ribbons treated with the spatial mosquito repellent, transfluthrin. Malar J. 2019; doi:10.1186/s12936-019-3048-8

44. Cohuet A, Simard F, Wondji CS, Antonio-nkondjio C, Awono-ambene P, Fontenille D. High Malaria Transmission Intensity Due to Anopheles funestus (Diptera_l_: Culicidae) in a Village of Savannah – Forest Transition Area in Cameroon. 2004; 901–905.

45. Trape JF, Tall A, Sokhna C, Ly AB, Diagne N, Ndiath O, et al. The rise and fall of malaria in a west African rural community, Dielmo, Senegal, from 1990 to 2012: A 22 year longitudinal study. Lancet Infect Dis. 2014;14. doi:10.1016/S1473-3099(14)70712-1

46. Antonio-Nkondjio C, Awono-Ambene P, Toto JC, Meunier JY, Zebaze-Kemleu S, Nyambam R, et al. High malaria transmission intensity in a village close to Yaounde, the capital city of Cameroon. J Med Entomol. 2002;39. doi:10.1603/0022-2585-39.2.350

47. Gleave K, Lissenden N, Richardson M, Choi L, Ranson H. Piperonyl butoxide (PBO) combined with pyrethroids in insecticide-treated nets to prevent malaria in Africa. Cochrane Database of Systematic Reviews. 2018. doi:10.1002/14651858.CD012776.pub2

48. Protopopoff N, Mosha JF, Lukole E, Charlwood JD, Wright A, Mwalimu CD, et al. Effectiveness of a long-lasting piperonyl butoxide-treated insecticidal net and indoor residual spray interventions, separately and together, against malaria transmitted by pyrethroid-resistant mosquitoes: a cluster, randomised controlled, two-by-two factorial design trial. Lancet. 2018;391. doi:10.1016/S0140-6736(18)30427-6

49. Pennetier C, Bouraima A, Chandre F, Piameu M, Etang J, Rossignol M, et al. Efficacy of Olyset® Plus, a New Long-Lasting Insecticidal Net Incorporating Permethrin and Piperonil-Butoxide against Multi-Resistant Malaria Vectors. PLoS One. 2013;8. doi:10.1371/journal.pone.0075134

50. Corbel V, Chabi J, Dabiré RK, Etang J, Nwane P, Pigeon O, et al. Field efficacy of a new mosaic long-lasting mosquito net (PermaNet® 3.0) against pyrethroid-resistant malaria vectors: A multi centre study in Western and Central Africa. Malar J. 2010;9. doi:10.1186/1475-2875-9-113

51. Ministry of Health and Social Welfare. Tanzania national Malaria Strategic Plan 2014-2020 [Internet]. Available: https://www.out.ac.tz/wp-content/uploads/2019/10/Malaria-Strategic-Plan-2015-2020-1.pdf

52. World Health Organization. Larval source management: a supplementary malaria vector control measure [Internet]. 2013. Available: https://apps.who.int/iris/handle/10665/85379

53. Mashauri FM, Manjurano A, Kinung’hi S, Martine J, Lyimo E, Kishamawe C, et al. Indoor residual spraying with microencapsulated pirimiphos-methyl (Actellic® 300CS) against malaria vectors in the Lake Victoria basin, Tanzania. PLoS One. 2017; doi:10.1371/journal.pone.0176982

54. Haji KA, Thawer NG, Khatib BO, Mcha JH, Rashid A, Ali AS, et al. Efficacy, persistence and vector susceptibility to pirimiphos-methyl (Actellic®300CS) insecticide for indoor residual spraying in Zanzibar. Parasites and Vectors. 2015;8. doi:10.1186/s13071-015-1239-x

